# Personalized Single-Cell Proteogenomics to Distinguish Acute Myeloid Leukemia from Non-Malignant Clonal Hematopoiesis

**DOI:** 10.1101/2020.10.22.20216028

**Authors:** Laura W. Dillon, Jack Ghannam, Chidera Nosiri, Gege Gui, Meghali Goswami, Katherine R. Calvo, Katherine E. Lindblad, Karolyn A. Oetjen, Matthew Wilkerson, Anthony R. Soltis, Gauthaman Sukumar, Clifton L. Dalgard, Julie Thompson, Janet Valdez, Christin B. DeStefano, Catherine Lai, Adam Sciambi, Robert Durruthy-Durruthy, Aaron Llanso, Saurabh Gulati, Shu Wang, Aik Ooi, Pradeep K. Dagur, J. Philip McCoy, Patrick Burr, Yuesheng Li, Christopher S. Hourigan

**Affiliations:** Laboratory of Myeloid Malignancies, Hematology Branch, National Heart, Lung, and Blood Institute, National Institutes of Health, Bethesda, MD 20892 United States; Department of Laboratory Medicine, Clinical Center, National Institutes of Health, Bethesda, MD 20892 United States; Precision Medicine Initiative for Military Medical Research and Education, Uniformed Services University of the Health Sciences, Bethesda, MD 20814; Henry M. Jackson Foundation for the Advancement of Military Medicine, Rockville, MD 20817; Department of Anatomy, Physiology and Genetics, Uniformed Services University of the Health Sciences, Bethesda, MD 20814; The American Genome Center, Uniformed Services University of the Health Sciences, Bethesda, MD 20814; Mission Bio, Inc., South San Francisco, CA, 94080, USA; Flow Cytometry Core, National Heart, Lung, and Blood Institute, National Institutes of Health, Bethesda, MD 20892 United States; DNA Sequencing and Genomics Core, National Heart, Lung, and Blood Institute, National Institutes of Health, Bethesda, MD 20892 United States

**Author notes:** **Corresponding author:** Christopher S. Hourigan, DM, DPhil, Laboratory of Myeloid Malignancies, National Heart, Lung, and Blood Institute, National Institutes of Health, Room 10CRC 5-5130, 10 Center Drive, Bethesda, Maryland 20814-1476. These authors contributed equally (Laura W. Dillon, Jack Ghannam, Chidera Nosiri).

## Abstract

Genetic mutations associated with acute myeloid leukemia can also be detected in age-related clonal hematopoiesis, making confident assignment of detected variants to malignancy challenging particularly in the post-treatment setting. This has implications for measurable residual disease monitoring, where the relationship between sequencing and flow cytometry is also imperfect. We show, using whole-genome-sequencing informed patient-personalized single-cell DNA and antibody-oligonucleotide sequencing, that it is possible to resolve immunophenotypic identity of clonal architecture.

## Main text

Age-related clonal hematopoiesis (also known as clonal hematopoiesis of indeterminate potential) is seen in those without hematological malignancy and with a somatic mutation detectable in a clonal population of mature blood cells^1-3^. These mutations, most commonly in epigenetic regulators *DNMT3A* and *TET2*, typically are found in older individuals and appear to be associated with an increased risk for death, predominately from inflammation-associated vascular events^4^. These mutations are also common in patients with Acute Myeloid Leukemia (AML)^5^ and can make genomic measurement of residual disease challenging^6,7^. Single-cell DNA-sequencing (scDNA-seq) for AML has recently been described^8,9^ but has been unable to detect novel chromosomal fusions that are the key, and often only, defining feature for multiple subtypes of this disease. We performed deep laboratory interrogation of samples, using whole-genome and targeted error-corrected sequencing, scDNA-seq with antibody-oligonucleotide conjugates and multiparameter flow cytometry, from three adult patients with relapsed AML who harbored mutations detected potentially attributable to either leukemia or clonal hematopoiesis (Figure 1A).

**Figure 1.**
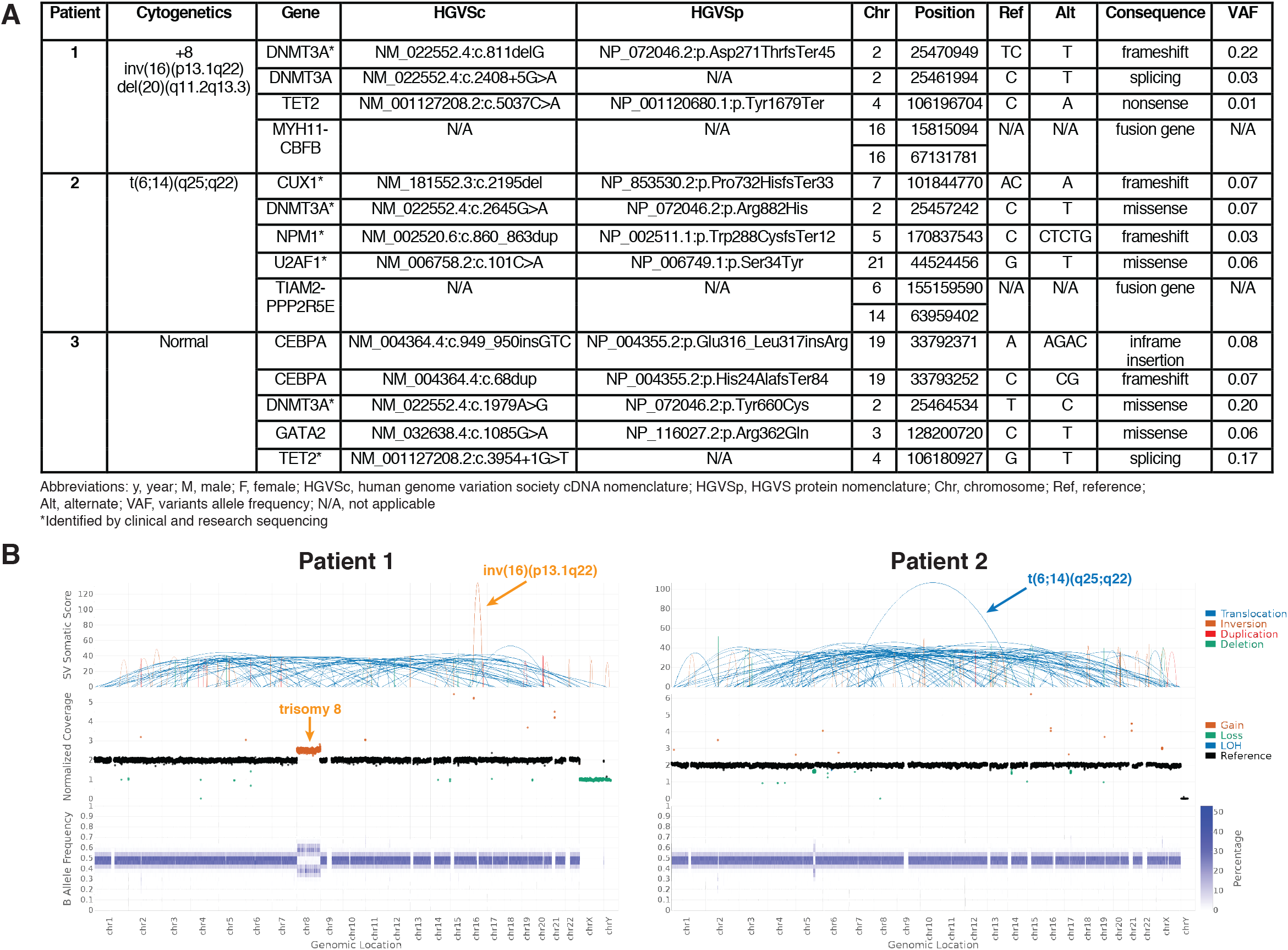
Clinical and genetic characterization. (A) Patient characteristics and AML genetics (B) Whole genome sequencing identified structural aberrations consistent with cytogenetic studies (chromosomal locations displayed on the x-axis). Top: structural changes identified by the structural variant caller. Middle: Total normalized somatic copy number estimates. Bottom: B-allele frequency at binned regions across the genome. Additional genetic material from chromosome 8 along with allelic imbalance was observed in patient 1, consistent with trisomy 8. Data visualized using Illumina Analysis Software.

Patient 1 was a man in his mid 70s with a second relapse of AML. Comorbidities included type 2 diabetes with neuropathy, retinopathy and peripheral vascular disease, hypertension, hyperlipidemia, coronary arterial disease, congestive heart failure, active tobacco use with chronic obstructive pulmonary disease and obstructive sleep apnea. Prior chemotherapy included clofarabine and cytarabine. Clinical flow cytometry identified a 1% CD34 positive population of abnormal blasts in blood. Blood marrow metaphase cytogenetics revealed a 47,XY karyotype with inversion of chromosome 16 and trisomy of chromosome 8 in four metaphases. A mutation in *DNMT3A* was noted by clinical DNA-sequencing, with additional mutations in *DNMT3A* and *TET2* also noted in error-corrected research DNA-sequencing.

Patient 2 was a woman in her mid 60s with a second relapse of AML. No vascular, diabetic, or other comorbidities were noted. Prior chemotherapy had included three rounds of intensive chemotherapy (idarubicin and cytarabine followed by cytarabine consolidation, then salvage therapy with cytarabine, etoposide and mitoxantrone). Clinical flow cytometry identified a 9% CD117 positive population of abnormal blasts in bone marrow. CD117 immunohistochemistry was positive for 10% of cells. Translocation t(6;14)(q25;q22) was observed in nine of twenty metaphases examined. Clinical DNA-sequencing noted mutations in *DNMT3A, U2AF1, NPM1* and *CUX1*.

Patient 3 was a man in his early 70s with a first relapse of AML. Concurrent comorbidities included hypertension and hyperlipidemia. He had previously received intensive chemotherapy with idarubicin and cytarabine induction followed by cytarabine consolidation. Clinical flow cytometry identified a 4.4% CD117 positive population of abnormal blasts in bone marrow. Clinical DNA-sequencing noted mutations in both *DMNT3A* and *TET2* and research sequencing confirmed these and also identified mutations in *GATA2* and *CEBPA*.

In all three patients the relationship between mutations discovered by targeted DNA-sequencing, leukemia-associated chromosomal abnormalities, and aberrant immunophenotype was unclear based on clinical evaluation.

In the first two patients, with known chromosomal structural abnormality from clinical metaphase cytogenetics, whole genome sequencing was performed for exact DNA breakpoint determination using bone marrow aspirate sorted for either CD34 or CD117 positivity based on clinical immunophenotype (Figure 1B, Supplementary Figure 1). Patient specific primers for scDNA-seq (Mission Bio, Inc.) were then designed to detect these breakpoints and also mutations identified by clinical and research targeted DNA-sequencing (Supplementary Table 1). Both chromosomal breakpoints and mutations found by bulk targeted DNA-sequencing were detected by scDNA-seq (Supplementary Figure 2).

Profiling cell surface protein expression using antibody-oligo conjugates together with scDNA-seq has recently been described^10^. The use of antibody-oligo conjugates in these scDNA-seq experiments could approximate the cell-surface immunophenotypic expression distributions as benchmarked by multiparameter flow cytometry (Supplementary Figure 3). This ability to pair cell surface immunophenotype with scDNA-seq genotyping for both chromosomal aberrations and sequence mutations allowed us to fully resolve the relationship between clonal hematopoiesis and AML in all three patients.

Five independent genetically-defined clones were found in peripheral blood mononuclear cells (PBMCs) of Patient 1 (Figure 2A). The pathognomonic inversion 16 breakpoint was detected in 13% of cells examined and was mutually exclusive from clones containing *DNMT3A* and *TET2* mutations. Trisomy of chromosome 8 was found in cells containing inversion 16 but not in other clones. While clinical flow cytometry described a 1% abnormal blast population positive for CD13, CD34 and CD117, this particular immunophenotype represented only half of the cells genotyped as containing inversion 16 and trisomy 8. The leukemic genotype was also found in cells with an immunophenotype consistent with more mature myeloid cell types (CD11b, CD33 and CD123 positive) but notably not in T (CD3) or B (CD19) lymphocytes. In contrast, the three subclones each containing one mutation in either *DNMT3A* or *TET2* did not have any restriction to myeloid or lymphoid lineages.

**Figure 2.**
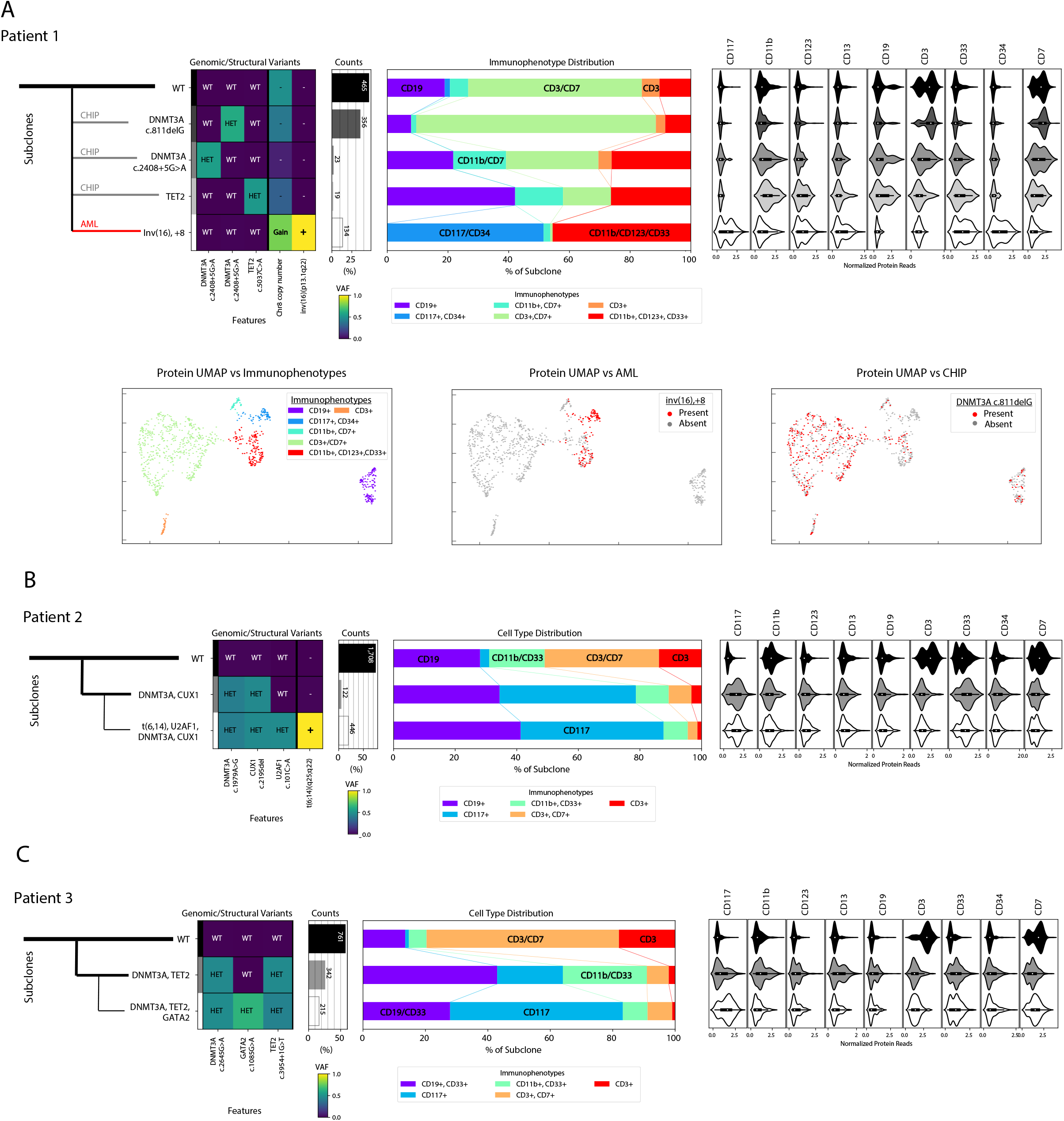
Independent and sequential development of leukemia from clonal hematopoiesis. Clonal architecture as determined by single-cell DNA and antibody-oligonucleotide sequencing. Left: Genomic subclones with wild type (WT), heterozygous (HET), present (Gain/+) or absent (-) features. Center: Immunophenotype as a percentage of each subclone. Right: Cell surface protein expression for each subclone. For patient 1, cells clustered by cell surface protein expression (2A lower), UMAP shown colored by immunophenotype (left) and genotype, with both leukemic subclone (middle) and one clonal hematopoiesis subclone (right) depicted.

Unlike this first case, patients 2 and 3 both had AML that likely arose from a preceding clonal hematopoiesis cell population. ScDNA-seq of bone marrow mononuclear cells (BMMCs) from patient 2 demonstrated that 20% of cells contained the leukemia defining features of t(6;14) translocation and mutated *U2AF1*, in addition to the *DNMT3A* and *CUX1* mutations which were within but not specific to this clone (Figure 2B). The *NPM1* mutation was incompletely genotyped. This malignant clone had a diverse range of immunophenotypes including both CD117 positive and negative populations. Cells containing mutations in *DNMT3A* and *CUX1* but not the t(6;14) translocation and *U2AF1* mutation represented just 5% of cells.

ScDNA-seq of BMMCs from patient 3 identified a clone representing 16% of cells which contained *DNMT3A, TET2, CEBPA* and *GATA2* mutations and was markedly enriched for CD117 cell surface protein expression (Figure 2C). *CEBPA* was incompletely genotyped but only found within cells with mutated *GATA2*. Cells containing mutations in *DNMT3A* and *TET2* but not *GATA2* or *CEBPA*, presumably representing the founder population of non-malignant clonal hematopoiesis, represented 26% of all bone marrow cells sequenced and had a diverse spectrum of immunophenotypes ranging across both myeloid and lymphoid lineages.

This is the first description of whole-genome sequencing informed patient-personalized scDNA-seq to accurately distinguish leukemic cells from clonal hematopoiesis. This is also the first direct comparison of antibody-oligonucleotide and scDNA-seq with concurrent multiparameter flow cytometry. The ability to determine the optimum combinations of immunophenotypic markers necessary to identify malignant subclones may have utility in flow cytometric approaches for measurement of residual AML, which currently underrepresent total genetically-defined leukemic burden. While targeted DNA-sequencing cannot determine if mutations associated with clonal hematopoiesis are also represented within the malignant clone, detection of pathognomonic fusion genes by single-cell DNA and antibody-oligonucleotide sequencing can resolve, and determine the immunophenotype of, the relevant leukemic clonal architecture allowing comprehensive and personalized assessment of AML. This integrated multimodal approach has implications for precision medicine in AML and other diseases.

## Methods

### Clinical samples

Bone marrow (BM) aspirate and peripheral blood (PB) samples were collected from patients with relapsed or refractory AML after receipt of written informed consent on a National Heart, Lung and Blood Institute institutional review board–approved protocol in accordance with the Declaration of Helsinki. BM mononuclear cells (BMMCs) were purified from BM aspirate and peripheral blood mononuclear cells (PBMCs) from PB using density centrifugation, cryopreserved, and stored in liquid nitrogen. Genomic DNA (gDNA) was isolated either from fresh heparinized BM aspirate using the QIAamp DNA Blood Midi kit or from thawed PBMCs or BMMCs using the QIAamp DNA Micro kit (Qiagen).

### Targeted DNA sequencing

Mutations were identified in each patient by error-corrected DNA-sequencing of 50ng of gDNA using a panel covering 75 genes commonly mutated in myeloid malignancies (Myeloid VariantPlex, ArcherDx). The resulting libraries were subjected to paired-end 150-bp sequencing on a Miseq (Illumina). Raw sequencing FASTQ files were analyzed using the Archer Analysis software version 6.0.2.3. AML-associated variants (Figure 1A), with a minimum variant allele frequency (VAF) of 0.01 were identified using information regarding UMI read families, background error rate models, unique start sites, strand-specific priming, homopolymer runs, and predicted variant consequence.

### Whole genome sequencing

For patients with known cytogenetic rearrangements (Patients 1 and 2), whole genome sequencing (WGS) was performed to determine the DNA breakpoint locations. gDNA isolated from buccal swabs and microbead (Miltenyi Biotech)-positively enriched CD34 or CD117 positive cells from BMMCs was used for WGS library preparation using the TruSeq DNA PCR-Free kit with ligation of IDT for Illumina TruSeq DNA UD Indexes. Libraries were clustered in a single library per lane topography with (5 lanes per AML library, 1 lane per germline library) on a cBot2 (Illumina) before sequencing on a HiSeq X platform (Illumina) to generate paired-end 150bp reads. Whole genome sequencing raw data was converted from BCL to FASTQ format using Illumina’s bcl2fastq2 v2.20. Samples were processed by the HAS2.2 (Illumina) resequencing analysis workflow. Reads were aligned to the hg38 reference genome plus decoy sequences using isaac (version Isaac-04.17.06.15). Then, matched tumor and normal results were analyzed by Illumina’s tumor normal workflow, which included somatic variant detection by strelka2 (version 2.80) and somatic structural variant detection by manta (version 1.1.1) and somatic copy number detection by canvas (version 1.28.0.272). Breakpoint locations for fusion partners was chosen for further analysis (Figure 1AB, Supplementary Figure 1).

### Single-cell DNA and Protein sequencing

Single-cell DNA sequencing with antibody-oligonucleotide staining was performed using the MissionBio Tapestri single-cell DNA sequencing V2 platform, per manufacturer’s instructions. In short, custom single-cell DNA sequencing panels targeting the patient-specific mutations and chromosomal structural abnormalities (Figure 1A, Supplementary Table 1) and oligonucleotide-conjugated antibodies (AOC) targeting cell surface proteins of interest (Supplementary Table 2) were designed and manufactured by MissionBio. Approximately 815,000 thawed BMMCs or PBMCs were stained for 30 min at room temperature with the AOC pool followed by four rounds of washing with DPBS containing 7% FBS. Stained cells were resuspended in cell buffer and subjected to microfluidic encapsulation, lysis, and cell barcoding on the Tapestri instrument. Amplification of the targeted DNA regions and antibody oligonucleotide tags was performed by incubating the barcoded DNA emulsions in a thermocycler as follows: 98°C for 6 min (4°C/sec); 11 cycles of 95°C for 30 sec, 72°C for 10 sec, 61°C for 3 min, 72°C for 20 sec (1°C/sec); 13 cycles of 95°C for 30 sec, 72°C for 10 sec, 48°C for 3 min, 72°C for 20 sec (1°C/sec); and 72°C for 6 min (4°C/sec). Emulsions were broken and DNA digested and purified with 0.7X SPRISelect reagent (Beckman Coulter). The beads were pelleted, and the supernatant retained for antibody library preparation, while the remaining beads were washed with 80% ethanol and the DNA targets eluted in nuclease-free water. The supernatant containing the antibody tags was incubated with a biotinylated capture oligo (/5Biosg/CGAGATGACTACGCTACTCATGG/3C6/, Integrated DNA Technologies) at 96°C for 5 min, followed by ice for 5 min, and recovered with streptavidin beads (Dynabead MyOne Streptavidin C1, Thermo Fisher). Indexed Illumina libraries were generated by amplifying DNA libraries with MissionBio V2 Index Primers and protein libraries bound to streptavidin beads with i5 and i7 index primers (Integrated DNA Technologies) in the thermocycler using the following program: 95°C for 3 min; 10 cycles (DNA library) or 20 cycles (protein library) of 98°C for 20 sec, 62°C for 20 sec, 72°C for 45 sec; 72°C for 2 min. Final libraries were purified with 0.69X (DNA) or 0.9X (protein) SPRISelect reagent. Libraries were pooled and subjected to paired-end 150-bp sequencing on a Novaseq 6000 (Illumina).

Raw FASTQ files were analyzed using the Tapestri pipeline (MissionBio) and custom python scripts. An average of 1,797 cells were sequenced per patient (Supplementary Table 3). SNVs and indels identified via GATK were filtered for quality score, read depth, and minimum frequency, after which UMAP dimension reduction and clustering were applied to identify subclone populations. The presence of inversions or translocations in those subclones were noted by counting amplicons spanning the splice junctions. Copy number variation was calculated by normalizing read counts for each DNA panel target to the wild type subpopulation and then observing a relative gain or loss of reads. Protein expression was calculated with a centered-log ratio transformation on the raw antibody counts to normalize for read depth variation between cells. From that, immunophenotypes were identified via UMAP dimension reduction and k-means clustering; the protein signature of each cluster was calculated by observing when the median expression for each target surpassed a fixed threshold across proteins.

### Flow Cytometry

Thawed PBMCs or BMMCs were resuspended in Cell Staining Buffer (BioLegend) and blocked with Human TruStain FcX Fc receptor blocking solution (BioLegend) for 10 min. Cells were stained with a 1:100 dilution of Zombie UV™ Fixable Viability dye (BioLegend) for 10 min and then incubated with 5μL of each antibody listed in Supplementary Table 4 for 20 min. All steps were performed at ambient temperatures. At least 100,000 events were acquired with a BD FACSymphony; data were analyzed with FlowJo Software version 10 (BD).

## Data Availability

Raw data is available upon request and will be submitted to dbGaP prior to formal publication.

## Acknowledgements

This work was supported by the Intramural Research Program of the National Heart, Lung, and Blood Institute (NHLBI) of the National Institutes of Health (NIH) and by the Department of Defense award: HU0001-18-2-0038 to CLD.

This work utilized the NHLBI Flow Cytometry Core, the NHLBI Sequencing and Genomics Core and the computational resources of the NIH HPC Biowulf cluster (http://hpc.nih.gov).

The opinions and assertions expressed herein are those of the author(s) and do not necessarily reflect the official policy or position of the Uniformed Services University or the Department of Defense, or the Henry M. Jackson Foundation for the Advancement of Military Medicine, Inc.

## References

1. Xie, M., et al. Age-related mutations associated with clonal hematopoietic expansion and malignancies. Nat Med 20, 1472–1478 (2014).

2. Busque, L., et al. Recurrent somatic TET2 mutations in normal elderly individuals with clonal hematopoiesis. Nat Genet 44, 1179–1181 (2012).

3. Young, A.L., Challen, G.A., Birmann, B.M. & Druley, T.E. Clonal haematopoiesis harbouring AML-associated mutations is ubiquitous in healthy adults. Nat Commun 7, 12484 (2016).

4. Jaiswal, S., et al. Clonal Hematopoiesis and Risk of Atherosclerotic Cardiovascular Disease. N Engl J Med 377, 111–121 (2017).

5. Tyner, J.W., et al. Functional genomic landscape of acute myeloid leukaemia. Nature 562, 526–531 (2018).

6. Jongen-Lavrencic, M., et al. Molecular Minimal Residual Disease in Acute Myeloid Leukemia. N Engl J Med 378, 1189–1199 (2018).

7. Hourigan, C.S., et al. Impact of Conditioning Intensity of Allogeneic Transplantation for Acute Myeloid Leukemia With Genomic Evidence of Residual Disease. J Clin Oncol 38, 1273–1283 (2020).

8. Miles, L.A., et al. Single cell mutational profiling delineates clonal trajectories in myeloid malignancies. bioRxiv (2020).

9. Morita, K., et al. Clonal Evolution of Acute Myeloid Leukemia Revealed by High-Throughput Single-Cell Genomics. bioRxiv (2020).

10. Demaree, B., et al. Joint profiling of proteins and DNA in single cells reveals extensive proteogenomic decoupling in leukemia. bioRxiv (2020).

