## Supplementary Appendix for "Personalized Single-Cell Proteogenomics to Distinguish Acute Myeloid Leukemia from Non-Malignant Clonal Hematopoiesis"

**Supplementary Table 1:** Single-Cell DNA Sequencing Custom Panel Amplicon Coverage

| Chromosome | Start Location | End Location | Target | Custom Panel |
|---|---|---|---|---|
| 1 | 115256295 | 115256570 | NRAS | CO101 |
| 1 | 115258524 | 115258799 | NRAS | CO101 |
| 1 | 115258610 | 115258825 | NRAS | CO86 |
| 2 | 25457051 | 25457294 | DNMT3A | CO86 |
| 2 | 25457113 | 25457372 | DNMT3A | CO101 |
| 2 | 25461785 | 25462020 | DNMT3A | CO101 |
| 2 | 25461795 | 25462064 | DNMT3A | CO86 |
| 2 | 25464503 | 25464762 | DNMT3A | CO86 |
| 2 | 25464512 | 25464762 | DNMT3A | CO101 |
| 2 | 25469541 | 25469741 | DNMT3A | CO101 |
| 2 | 25469542 | 25469741 | DNMT3A | CO86 |
| 2 | 25469874 | 25470130 | DNMT3A | CO101 |
| 2 | 25469875 | 25470130 | DNMT3A | CO86 |
| 2 | 25470295 | 25470550 | DNMT3A | CO101 |
| 2 | 25470907 | 25471166 | DNMT3A | CO86 |
| 2 | 25470928 | 25471182 | DNMT3A | CO101 |
| 2 | 198266732 | 198267007 | SF3B1 | CO101 |
| 2 | 198267133 | 198267406 | SF3B1 | CO101 |
| 2 | 209112874 | 209113149 | IDH1 | CO101 |
| 3 | 128200666 | 128200925 | GATA2 | CO86 |
| 3 | 128200674 | 128200934 | GATA2 | CO101 |
| 3 | 128202703 | 128202911 | GATA2 | CO101 |
| 3 | 128204886 | 128205082 | GATA2 | CO101 |
| 3 | 128204887 | 128205082 | GATA2 | CO86 |
| 3 | 128205085 | 128205300 | GATA2 | CO101 |
| 3 | 128205086 | 128205300 | GATA2 | CO86 |
| 4 | 55589584 | 55589859 | KIT | CO101 |
| 4 | 55599203 | 55599478 | KIT | CO101 |
| 4 | 106180769 | 106181018 | TET2 | CO86 |
| 4 | 106180826 | 106181059 | TET2 | CO101 |
| 4 | 106190625 | 106190861 | TET2 | CO101 |
| 4 | 106190626 | 106190861 | TET2 | CO86 |
| 4 | 106196565 | 106196781 | TET2 | CO101 |
| 4 | 106196566 | 106196781 | TET2 | CO86 |
| 5 | 113993585 | 113993800 | copy number | CO86 |
| 5 | 136633179 | 136633418 | copy number | CO86 |

| | | | | |
|---|---|---|---|---|
| 5 | 170837384 | 170837659 | NPM1 | CO101 |
| 5 | 170837463 | 170837704 | NPM1 | CO86 |
| 6 | 17076719 | 17076969 | copy number | CO101 |
| 6 | 40116142 | 40116388 | copy number | CO101 |
| 6 | 62094165 | 62094411 | copy number | CO101 |
| 7 | 4310243 | 4310467 | copy number | CO101 |
| 7 | 4310284 | 4310507 | copy number | CO86 |
| 7 | 4456902 | 4457130 | copy number | CO101 |
| 7 | 24149103 | 24149342 | copy number | CO101 |
| 7 | 101844541 | 101844801 | CUX1 | CO101 |
| 7 | 101844542 | 101844801 | CUX1 | CO86 |
| 7 | 101891713 | 101891983 | CUX1 | CO101 |
| 7 | 101891714 | 101891983 | CUX1 | CO86 |
| 7 | 101916615 | 101916874 | CUX1 | CO101 |
| 7 | 101916616 | 101916874 | CUX1 | CO86 |
| 7 | 105679464 | 105679690 | copy number | CO86 |
| 7 | 105679486 | 105679722 | copy number | CO101 |
| 7 | 124392444 | 124392651 | copy number | CO101 |
| 7 | 124392445 | 124392651 | copy number | CO86 |
| 7 | 137029771 | 137030010 | copy number | CO101 |
| 7 | 137029772 | 137030010 | copy number | CO86 |
| 7 | 148504626 | 148504901 | EZH2 | CO101 |
| 7 | 148506302 | 148506577 | EZH2 | CO101 |
| 7 | 148510911 | 148511147 | EZH2 | CO101 |
| 7 | 148510912 | 148511147 | EZH2 | CO86 |
| 7 | 148514326 | 148514561 | EZH2 | CO101 |
| 7 | 148514327 | 148514561 | EZH2 | CO86 |
| 8 | 28410869 | 28411109 | copy number | CO101 |
| 8 | 28410870 | 28411109 | copy number | CO86 |
| 8 | 144656722 | 144656981 | copy number | CO86 |
| 8 | 144656723 | 144656983 | copy number | CO101 |
| 9 | 5073540 | 5073815 | JAK2 | CO101 |
| 10 | 5554170 | 5554419 | copy number | CO101 |
| 10 | 77210063 | 77210313 | copy number | CO101 |
| 10 | 106721486 | 106721736 | copy number | CO101 |
| 11 | 32413388 | 32413663 | WT1 | CO101 |
| 11 | 32414173 | 32414432 | WT1 | CO101 |
| 11 | 32417743 | 32418018 | WT1 | CO101 |
| 11 | 32417847 | 32418088 | WT1 | CO86 |

| | | | | |
|---|---|---|---|---|
| 12 | 25380237 | 25380490 | KRAS | CO101 |
| 12 | 25398160 | 25398435 | KRAS | CO101 |
| 12 | 25398227 | 25398433 | KRAS | CO101 |
| 12 | 112888094 | 112888351 | PTPN11 | CO101 |
| 12 | 112888115 | 112888350 | PTPN11 | CO101 |
| 12 | 112926826 | 112927063 | PTPN11 | CO101 |
| 13 | 28592472 | 28592747 | FLT3 | CO101 |
| 13 | 28602154 | 28602429 | FLT3 | CO101 |
| 13 | 28607996 | 28608176 | FLT3 | CO101 |
| 13 | 28608167 | 28608392 | FLT3 | CO101 |
| 13 | 28609520 | 28609795 | FLT3 | CO101 |
| 14 | 56968883 | 56969129 | copy number | CO101 |
| 15 | 90631737 | 90632009 | IDH2 | CO101 |
| 16 | 8569694 | 8569944 | copy number | CO101 |
| 16 | 55770511 | 55770757 | copy number | CO101 |
| 17 | 7576929 | 7577204 | TP53 | CO101 |
| 17 | 7577048 | 7577317 | TP53 | CO86 |
| 17 | 7577375 | 7577637 | TP53 | CO101 |
| 17 | 7578061 | 7578319 | TP53 | CO101 |
| 17 | 7578362 | 7578627 | TP53 | CO101 |
| 17 | 7578364 | 7578623 | TP53 | CO86 |
| 17 | 74732864 | 74733069 | SRSF2 | CO101 |
| 17 | 74732876 | 74733145 | SRSF2 | CO86 |
| 18 | 9750542 | 9750791 | copy number | CO101 |
| 18 | 42531846 | 42532088 | SETBP1 | CO101 |
| 18 | 42531847 | 42532088 | SETBP1 | CO86 |
| 19 | 33792251 | 33792503 | CEBPA | CO101 |
| 19 | 33792252 | 33792503 | CEBPA | CO86 |
| 19 | 33793075 | 33793344 | CEBPA | CO86 |
| 19 | 33793075 | 33793337 | CEBPA | CO101 |
| 20 | 31021139 | 31021365 | ASXL1 | CO101 |
| 20 | 31021140 | 31021365 | ASXL1 | CO86 |
| 20 | 31022226 | 31022496 | ASXL1 | CO86 |
| 20 | 31022347 | 31022608 | ASXL1 | CO101 |
| 20 | 31022879 | 31023130 | ASXL1 | CO101 |
| 20 | 39486899 | 39487140 | copy number | CO101 |
| 20 | 39486982 | 39487223 | copy number | CO86 |
| 20 | 51296130 | 51296375 | copy number | CO101 |
| 20 | 51296131 | 51296375 | copy number | CO86 |

| | | | | |
|---|---|---|---|---|
| 21 | 36164767 | 36165018 | RUNX1 | CO101 |
| 21 | 36171457 | 36171732 | RUNX1 | CO101 |
| 21 | 36206683 | 36206907 | RUNX1 | CO101 |
| 21 | 36231582 | 36231857 | RUNX1 | CO101 |
| 21 | 36252788 | 36253030 | RUNX1 | CO101 |
| 21 | 36252793 | 36253028 | RUNX1 | CO86 |
| 21 | 36259140 | 36259430 | RUNX1 | CO101 |
| 21 | 36259141 | 36259430 | RUNX1 | CO86 |
| 21 | 44514678 | 44514947 | U2AF1 | CO101 |
| 21 | 44524257 | 44524532 | U2AF1 | CO101 |
| 21 | 44524417 | 44524634 | U2AF1 | CO86 |
| 22 | 33559405 | 33559641 | copy number | CO101 |
| X | 123191710 | 123191949 | STAG2 | CO101 |
| X | 123191711 | 123191949 | STAG2 | CO86 |
| X | 123200141 | 123200401 | STAG2 | CO101 |
| X | 123200142 | 123200401 | STAG2 | CO86 |
| X | 123220359 | 123220602 | STAG2 | CO101 |
| X | 123220360 | 123220602 | STAG2 | CO86 |
| X | 133549065 | 133549325 | PHF6 | CO101 |
| X | 133549066 | 133549325 | PHF6 | CO86 |
| 16 | 67131781 | 67131888 | MYH11-CBFB | CO86,CO101 |
| 16 | 15815094 | 15815226 | MYH11-CBFB | CO86,CO101 |
| 6 | 155159463 | 155159589 | TIAM2-PPP2R5E | CO86,CO101 |
| 14 | 63959401 | 63959485 | TIAM2-PPP2R5E | CO86,CO101 |

*coordinates in Genome Reference Consortium Human Build 27 (hg19)

**Supplementary Table 2:** Antibodies Used for Single-Cell Surface Protein Sequencing

| Marker | Clone |
|---|---|
| CD117 | 104D2 |
| CD11b | M1/70 |
| CD123 | 6H6 |
| CD13 | WM15 |
| CD19 | HIB19 |
| CD3 | HIT3a |
| CD33 | P67.6 |
| CD34 | AC136 |
| CD38 | REA572 |
| CD45 | HI30 |
| CD7 | CD7-6B7 |
| CD90 | AF-9 |
| HLA-A,B,C | W6/32 |
| HLA-DR | L243 |
| HLA-DR,DP,DQ | Tu39 |

**Supplementary Table 3:** Single-Cell DNA Sequencing Performance Characteristics

| Patient | Panel | Read Pairs | Number of Cells | Average Reads per Cell | Percentage of Reads Assigned to Cells |
|---|---|---|---|---|---|
| 1 | CO101 | 86,830,744 | 1,161 | 40,304 | 53.89 |
| 2 | CO86 | 28,546,762 | 2,647 | 6,912 | 64.09 |
| 3 | CO86 | 22,630,471 | 1,584 | 8,831 | 61.81 |

**Supplementary Table 4.** Antibodies Used for Multiparametric Flow Cytometry

| Marker | Fluorochrome | Clone | Catalog # | Manufacturer |
|---|---|---|---|---|
| LIVE - DEAD | BUV395 | N/A | 423108 | BioLegend |
| CD117 | APC | 104D2 | 313206 | BioLegend |
| CD11B | FITC | ICRF44 | 301330 | BioLegend |
| CD123 | BV421 | 6H6 | 306018 | BioLegend |
| CD13 | APC/Cy7 | WM15 | 301710 | BioLegend |
| CD19 | PECF594 | HIB19 | 302252 | BioLegend |
| CD3 | BUV737 | SK7 | 612752 | BD Biosciences |
| CD33 | BV605 | P67.6 | 366612 | BioLegend |
| CD34 | Percp/Cy5.5 | 581 | 343522 | BioLegend |
| CD38 | BV510 | HIT2 | 303540 | BioLegend |
| CD45 | BV785 | H130 | 304048 | BioLegend |
| CD7 | PE/Cy7 | CD7-6B7 | 343114 | BioLegend |
| CD90 (Thy1) | PE | 5E 10 | 328110 | BioLegend |
| HLA-DR | BV650 | L243 | 307650 | BioLegend |

**Supplementary Figure 1. DNA breakpoint mapping of chromosomal structural abnormalities from whole genome sequencing.** Whole genome sequencing reads from (A) CD-34+ bone marrow from patient 1 and (B) CD-117+ bone marrow from patient 2 identified by the somatic structural variant detection algorithm to align to known cytogenetic rearrangements, inv(16)(p13.1q22) and t(6;14)(q25;q22), respectively, are shown using the integrated genome viewer (IGV) version 2.5.2. Location of the DNA breakpoints are indicated by a dashed line and genomic coordinates (hg19) indicated above. The DNA sequence and breakpoint coordinates of each fusion product are depicted below. The fusion product used for tracking by a custom single-cell DNA sequencing panel are underlined.

**A**

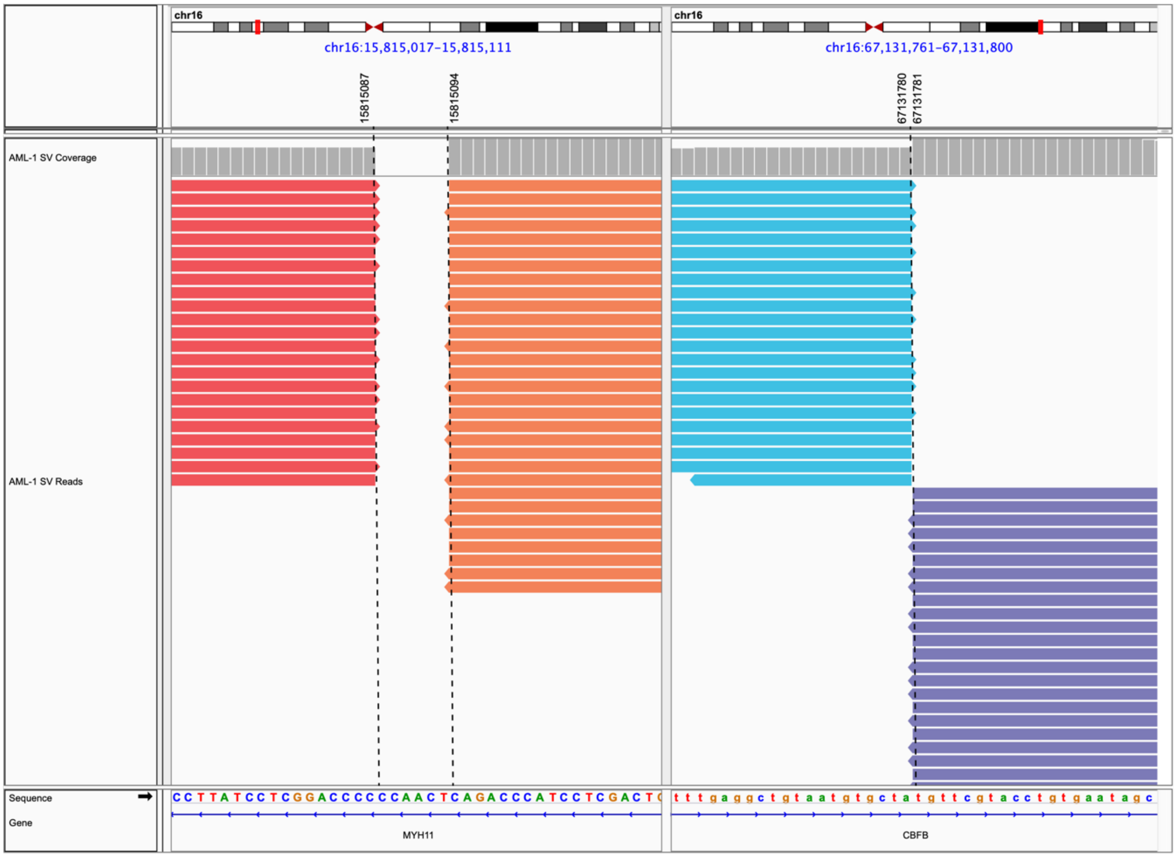


chr16:15815094::chr16:67131781

<u>**MYH11-CBFB**</u> GGCTGGACTGCAGCGGCTATTCACAGGTACGAACACAGACCCATCCTCGACTGCCATTCTCAGCCCCTCC

chr16:671131780::chr16:15815087

**CBFB-MYH11** TTGGTTTAGTCCAGGAATTTGAGGCTGTAATGTGCTAGGGGTCCGAGGATAAGGCTGGGGTCTGAACTTCT

**B**

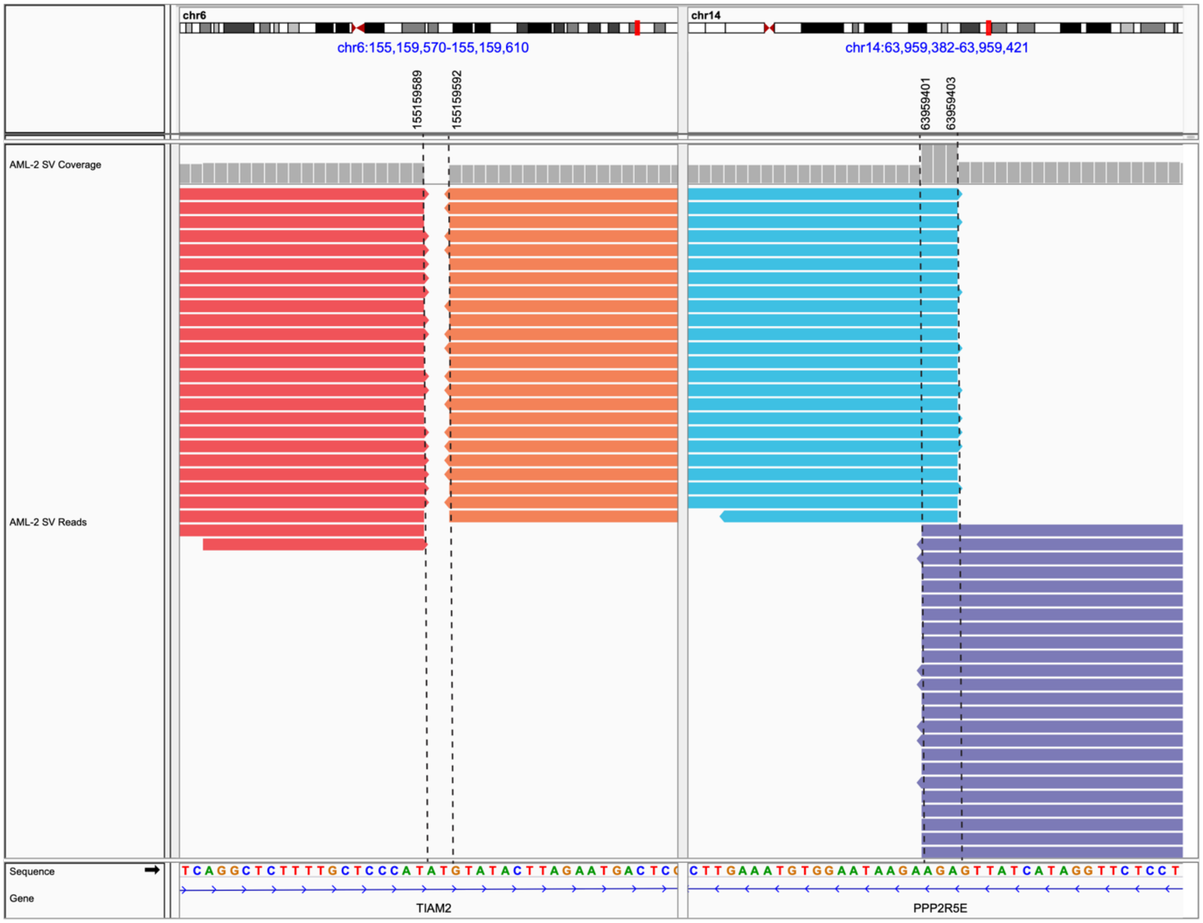


**chr6:155159589::chr14:63959401**

**TIAM2-PPP2R5E** **TTCCTCCTTTTTCATCAGGCTCTTTTGCTCCCATAGAGTTATCATAGGTTCTCCTTGATCATCAAGTG**

**chr1463959403::chr6:155159592**

**PPP2R5E-TIAM2** **CAAGTACAAACTTGAAATGTGGAATAAGAAGAGTATACTTAGAATGACTCGGGCTTCTTGTTCTAAA**

**Supplementary Figure 2. Comparison of variant allele frequency from bulk DNA sequencing and single-cell DNA sequencing.** The variant allele frequency (VAF) of each AML-associated variant (Figure 1A) interrogated by targeted error-corrected DNA sequencing performed on bulk DNA was plotted on the x-axis and by single-cell DNA sequencing on the y-axis. The Pearson correlation was calculated. Each dot represents one variant from one patient, with red, blue and green representing values from Patient 1, 2 and 3 respectively. A value of equivalence line is plotted as a dashed line. Prism version 8.4.3 was used for statistics and graphing.

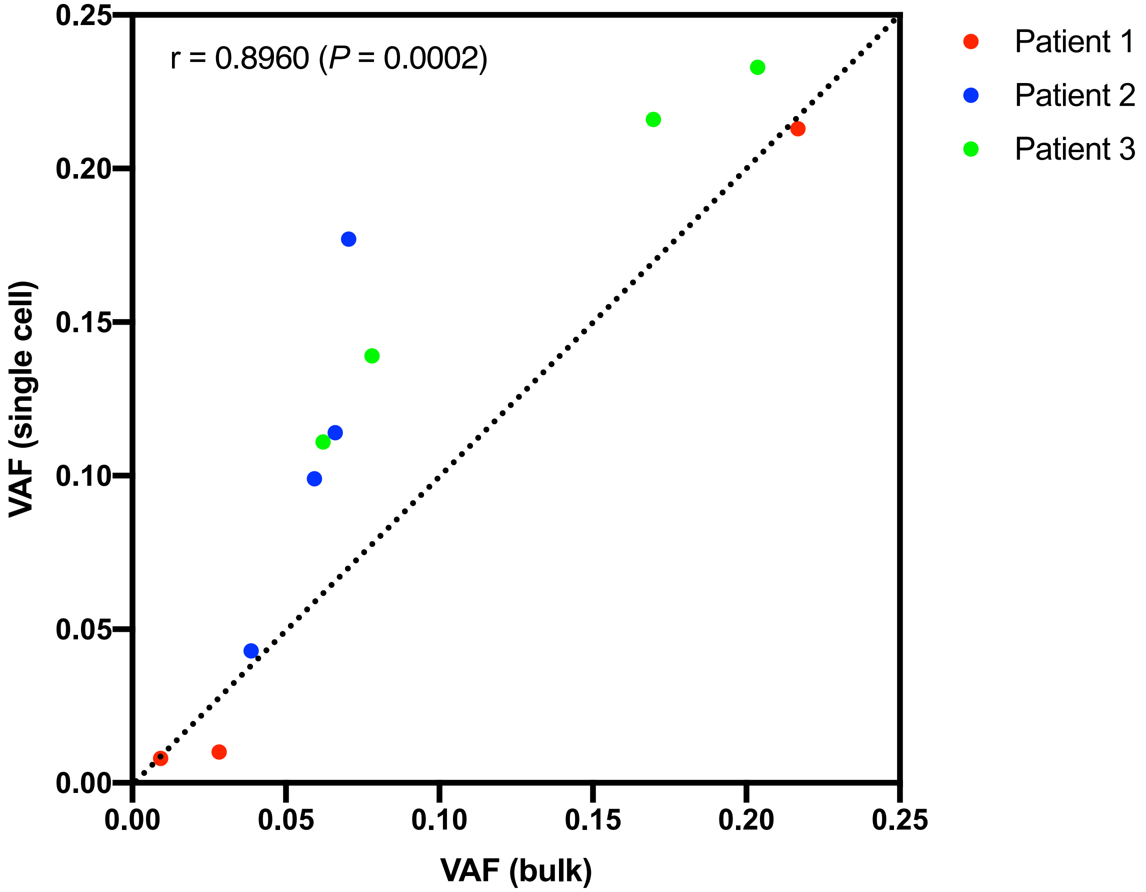

**Supplementary Figure 3. Comparison of antibodies from flow cytometry and single-cell surface protein expression.** The percentages of cells with positive antibody expression were compared for flow cytometry and single-cell sequencing. Marker counts <5 in flow or <10 in single-cell sequencing were considered as background and the data were log-transformed. Local Polynomial Regression (Loess) was implemented to obtain the optimal cutoffs for positive cells. The derivatives of predicted values were calculated to find the changing points of the distribution and the median was chosen if multiple points were detected. The percentages of positive cells were summarized and the Pearson correlation between the percentages from flow cytometry and single-cell sequencing data was calculated and plotted. Each dot represents one protein from one patient, with red, blue and green representing values from Patient 1, 2 and 3 respectively. A value of equivalence line is plotted as a dashed line. R 4.0.0 was used for model fitting, data processing and visualization.

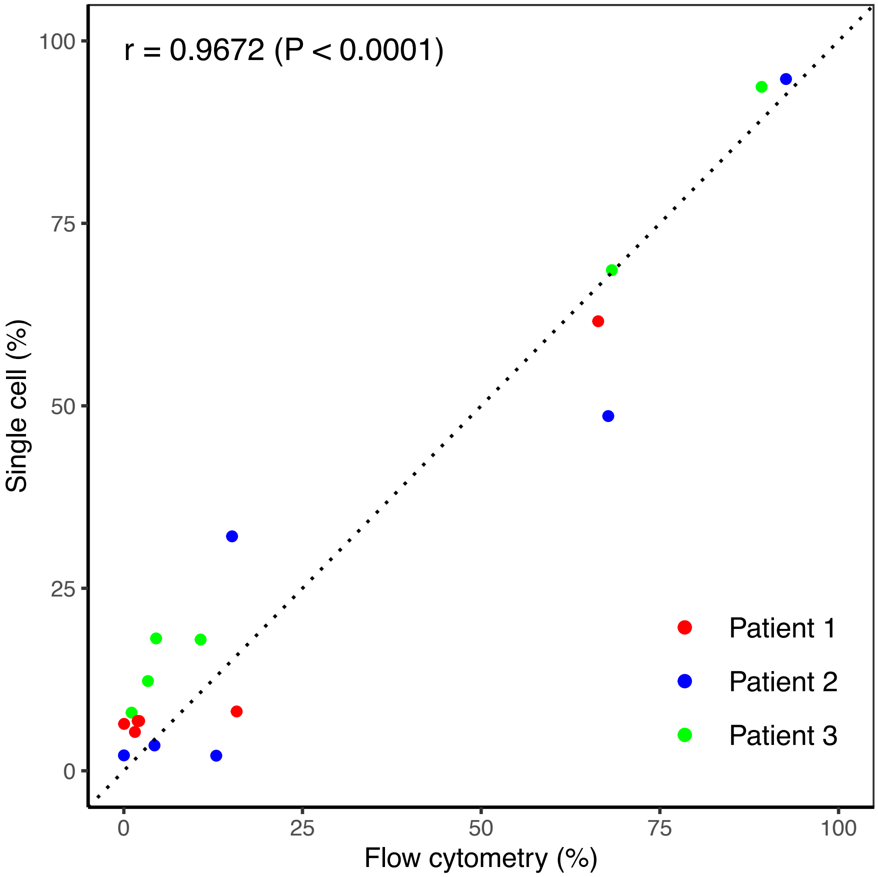
